# Early Evidence of Effectiveness of Digital Contact Tracing for SARS-CoV-2 in Switzerland

**DOI:** 10.1101/2020.09.07.20189274

**Authors:** Marcel Salathé, Christian L. Althaus, Nanina Anderegg, Daniele Antonioli, Tala Ballouz, Edouard Bugnion, Srdjan Čapkun, Dennis Jackson, Sang-Il Kim, James R. Larus, Nicola Low, Wouter Lueks, Dominik Menges, Cédric Moullet, Mathias Payer, Julien Riou, Theresa Stadler, Carmela Troncoso, Effy Vayena, Viktor von Wyl

**Author notes:** All authors contributed equally.

## Abstract

In the wake of the pandemic of coronavirus disease 2019 (COVID-19), contact tracing has become a key element of strategies to control the spread of severe acute respiratory syndrome coronavirus 2019 (SARS-CoV-2). Given the rapid and intense spread of SARS-CoV-2, digital contact tracing has emerged as a potential complementary tool to support containment and mitigation efforts. Early modelling studies highlighted the potential of digital contact tracing to break transmission chains, and Google and Apple subsequently developed the Exposure Notification (EN) framework, making it available to the vast majority of smartphones. A growing number of governments have launched or announced EN-based contact tracing apps, but their effectiveness remains unknown. Here, we report early findings of the digital contact tracing app deployment in Switzerland. We demonstrate proof-of-principle that digital contact tracing reaches exposed contacts, who then test positive for SARS-CoV-2. This indicates that digital contact tracing is an effective complementary tool for controlling the spread of SARS-CoV-2. Continued technical improvement and international compatibility can further increase the efficacy, particularly also across country borders.

---

Contact tracing is a key element of the response to the pandemic of coronavirus disease 2019 (COVID-19), caused by severe acute respiratory syndrome coronavirus 2019 (SARS-CoV-2). By August 30, 2020, nearly 25 million diagnosed cases and over 800,000 confirmed deaths had been recorded [1]. Contact tracing is part of a strategy of “test, trace, isolate and quarantine” (TTIQ), which aims to stop recently infected individuals from transmitting SARS-CoV-2 [2]. The capacity for transmission before the onset of symptoms and the short incubation period mean that contact tracing has to be fast to be effective [3,4]. While modelling results showed that rapid, digital contact tracing could be a critical tool for containment and mitigation efforts [5], early efforts to develop and deploy digital applications were hampered by critical limitations imposed by smartphone operating systems and concerns about confidentiality and data protection. The Exposure Notification (EN) framework, jointly developed by Google and Apple, addressed these limitations and enabled the implementation of digital contact tracing applications (apps) in which all proximity contact information, and any decision-making about whether or not to notify a user of an exposure, remain on a user’s device, rather than on the server of a central authority. This approach minimizes privacy risks [6], but the restriction of information to users’ devices means that data needed to evaluate the effectiveness of EN-based contact tracing apps have to be collected from different sources [7].

SwissCovid was the first EN-based contact tracing app launched by a governmental public health authority, initially as a pilot to a limited number of users, on May 25, 2020. On June 25, 2020 the application was made available to the general population. A growing number of governments have launched or announced EN-based contact tracing apps, but their effectiveness remains unknown. The significance of evaluating the effectiveness of EN-based contact tracing systems was highlighted by the World Health Organization [8] and is considered an ethical requirement for the continued deployment of such systems [9]. Here, we report early findings of the SwissCovid deployment from July 23, 2020 to September 10, 2020 (hereafter referred to as the study period).

## How does SwissCovid work?

The technical details of the SwissCovid system are given in [10], but briefly, SwissCovid uses the EN framework (version 1.2) to estimate proximity between phones. Each phone generates a daily Temporary Exposure Key (TEK), from which fast rotating proximity identifiers (RPI) are derived and exchanged with neighboring phones via Bluetooth Low Energy (BLE) beacons. Upon confirmation of SARS-CoV-2 infection by reverse transcriptase polymerase chain reaction (RT-PCR) testing, authorized Swiss health professionals can generate a single-use validation code (Covidcode), which is provided to the user. The Covidcode is associated with the beginning of the contagious period, which was determined to start 2 days before onset of symptoms [11] for symptomatic patients, and on the day of the test for patients who are asymptomatic at the time of testing. Upon entering a valid Covidcode in the app, TEKs for the contagious period are transmitted from the user’s phone to the computer server of the Swiss Federal Office of Public Health (FOPH). All SwissCovid apps regularly contact the server for uploaded TEKs and associated data, and compute the exposure risk for the previous 10 days. To ensure privacy, notifications are shown only on the phone and are not forwarded to a server. During the study period, the notification provided users with the last day of exposure, a reminder that they are entitled to a free RT-PCR test, and directed them to a SwissCovid-specific hotline (number only shown upon notification).

The BLE beacons, used by SwissCovid to estimate whether two devices have been in close proximity to each other for a period of time, are radio signals, which attenuate with distance. The EN framework application programming interface (API) estimates the amount of time a device has been close enough to other devices of infected individuals, based on three different attenuation intervals. To identify attenuation levels that would best estimate proximity below 1.5 meters, attenuation levels for different controlled and real-life scenarios produced by different devices at different distances and orientation were measured, and subsequently corrected using the per-device calibration values provided by the EN framework. Per-device calibration values take into account varying antenna and chip characteristics that influence sending and receiving powers. These results informed the parameterization of the EN framework so that SwissCovid users are notified if they spent at least 15 minutes in close proximity (1.5 meters) to RT-PCR-confirmed cases.

## Evaluation of privacy-preserving EN-based digital contact tracing

Digital contract tracing is a new method, and it is therefore crucial to rapidly evaluate and continuously monitor its effectiveness in the field. Digital contact tracing apps aim to prevent secondary transmissions by warning exposed contacts as early as possible [7]. To be an effective intervention, several technical, behavioral and procedural conditions must be met [12] (Figure 1). The app must be used by both the infected index case and exposed contacts (Figure 1, row 1), the index case must enter the Covidcode following a positive RT-PCR test (row 2), the exposed contacts must receive notifications (row 3), and the exposed contacts must respond to the warning in a timely fashion to prevent further transmission (rows 4 and 5). These conditions cannot be assessed directly via the app, due to the voluntary and decentralized nature of the system. Alternative indicators and data sources have therefore been developed or commissioned. In Switzerland, the Federal Statistical Office (FSO) monitors downloads and active use of SwissCovid and publicly releases the relevant number on a daily basis [13,14]. The FOPH updated the clinical registration form for RT-PCR-confirmed cases before the beginning of the study period, and included the SwissCovid app as an option for the reason for the test. In addition, several ongoing research studies collect information about app usage and notifications received on a monthly basis, e.g. Corona Immunitas [15], a nationwide Sars-CoV-2 seroprevalence study with digital follow-up surveys [16], and the COVID-19 Social Monitor (https://csm.netlify.app), a regular, longitudinal online-survey on social, economic, and behavioral aspects related to COVID-19, drawn from a representative panel for Switzerland. Furthermore, an ongoing cohort study embedded in contact tracing in the canton of Zurich [17], which enrolls RT-PCR-confirmed cases and their close contacts, investigates circumstances of transmissions and risk exposures to SARS-CoV-2, and collects data about use of the app.

**Figure 1:**
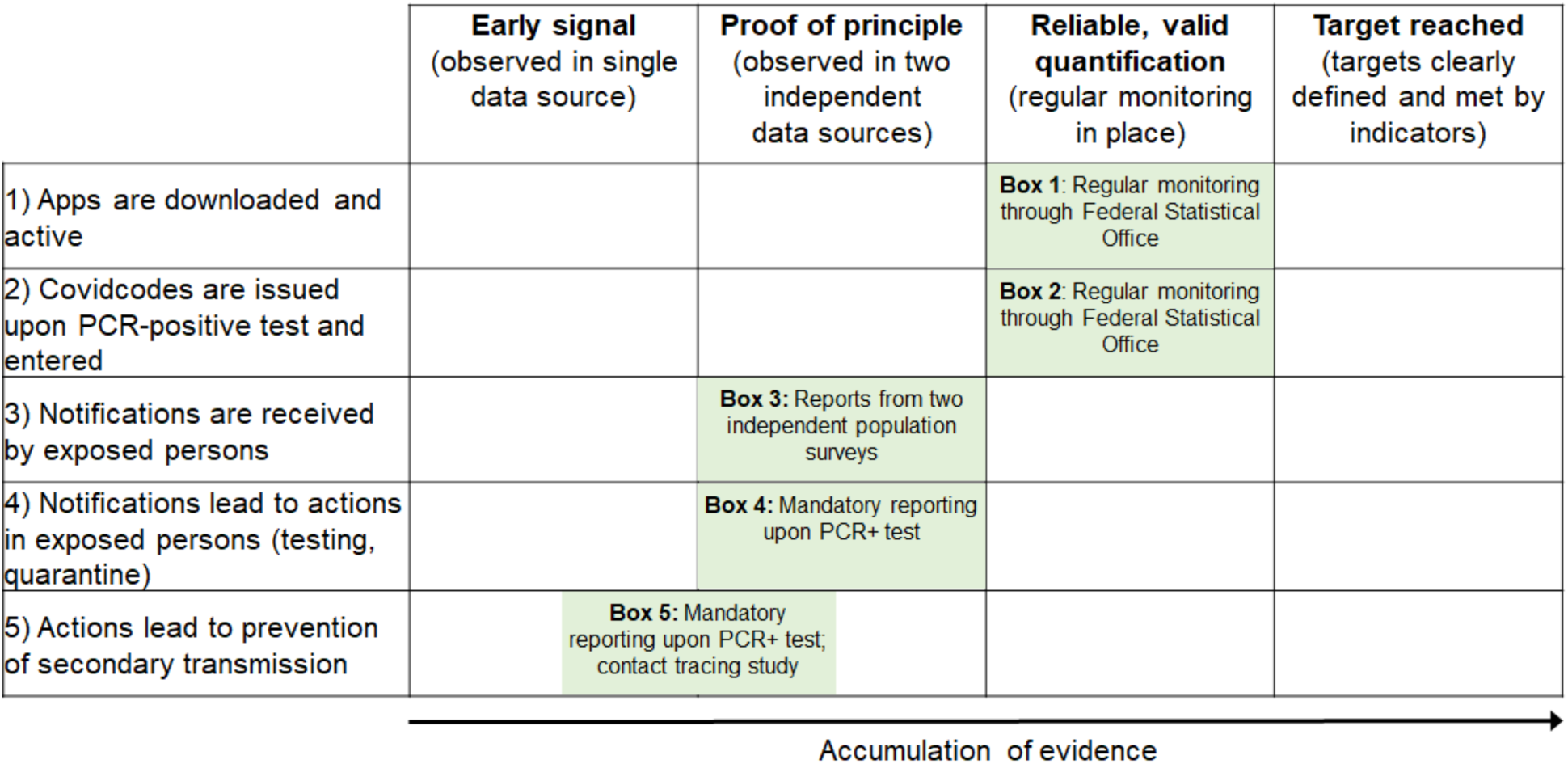
Quality of evidence for SwissCovid app effectiveness. Row labels: Necessary technical, procedural and behavioral conditions for digital contact tracing to be effective. Columns: Milestones in evidence accumulation. Green boxes indicate available data sources for evidence assessments; quantitative information related to green boxes are provided in the main text.

## Downloads and active use of SwissCovid app

To measure the number of app downloads and active apps, reliable monitoring is already in place (Figure 1, Box 1). By September 10, 2020, the SwissCovid app has been downloaded 2.36 million times, and the number of active apps per day has been estimated at 1.62 million (Figure 2; Figure 3A). The number of active users corresponds to 18.9% of the Swiss population (8.6 million).

**Figure 2:**
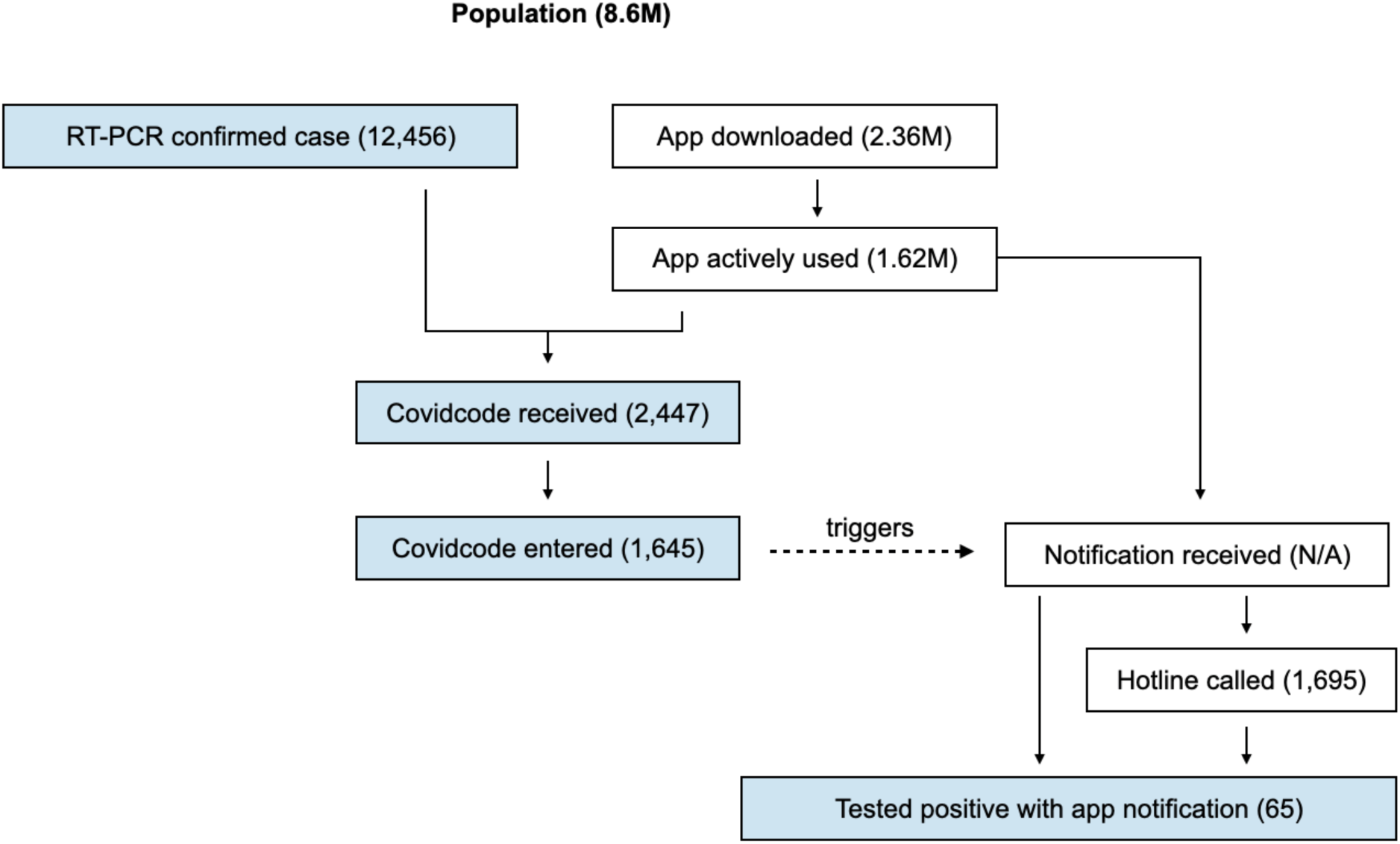
Flowchart of SwissCovid methodology. A person must have had a positive RT-PCR test and used the app in order to receive and enter a Covidcode that can trigger an exposure notification in other phones. Notifications are triggered by the decentralized mechanism described in the main text. The numbers shown are for the study period of July 23 to September 10, 2020. Blue box shading indicates positively tested individuals.

**Figure 3:**
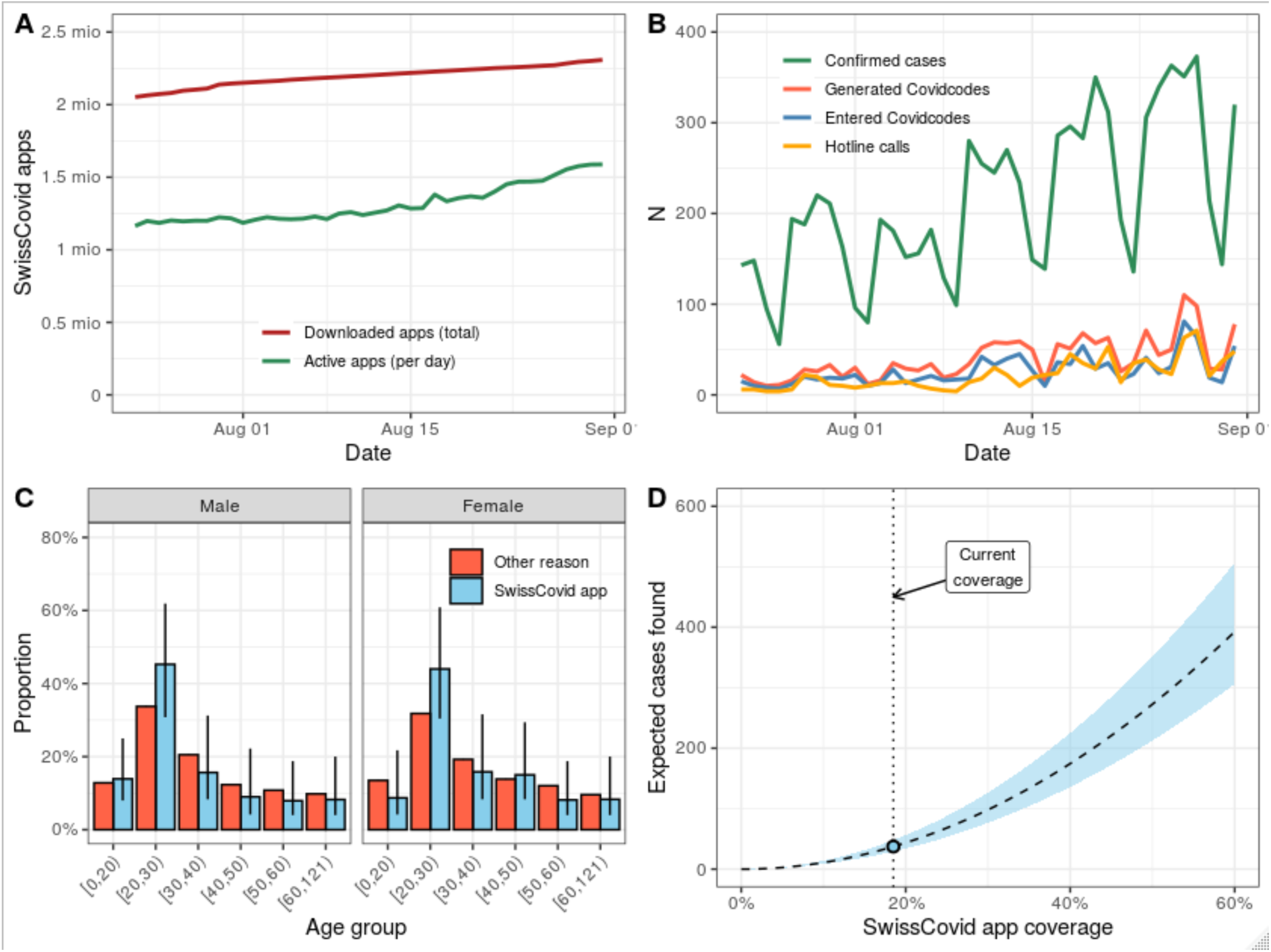
SwissCovid app measures. (A) Total number of downloaded apps and daily number of active apps. (B) Daily numbers of confirmed SARS-CoV-2 cases, generated Covidcodes, entered Covidcodes and hotline calls. (C) Age distribution of cases stratified by the reason for RT-PCR test (either SwissCovid app or other reason). (D) Expected number of RT-PCR-confirmed cases that were tested because of a notification by the app as a function of hypothetical app coverage during the study period. Error bars and the blue shaded area correspond to 95% confidence intervals.

During the study period, the FOPH reported 12,456 confirmed SARS-CoV-2 cases. During the same time period, the FOPH issued 2,447 (19.6% of confirmed cases) Covidcodes and 1,645 (13.2% of confirmed cases, 67.2% of issued Covidcodes) of these Covidcodes were entered in the app by the users (Figure 1, Box 2; Figure 3B). While the decentralized nature makes it impossible to know how many notifications were subsequently generated, the entered Covidcodes triggered 1,695 phone calls to the SwissCovid hotline, thus providing evidence for actions undertaken by notified contacts (Figure 1, Box 3; Figure 3B).

The Zurich SARS-CoV-2 Cohort is a longitudinal study embedded in the contact tracing of the canton of Zurich with continuous enrollment of RT-PCR-positive index cases and exposed contacts. Recruitment started on Aug. 7, and until Sept. 11, there were 235 index cases (median age 34 years, 51% males), and 185 (median age 33 years, 53% males) exposed contacts enrolled. Of the 235 index cases, 148 (63%) used the app. Of those 148 app users, 134 (91%) received, and 127 (86%) uploaded the Covidcode. Four cases did not upload the Covidcode because they had already informed their contacts when they received the code. Three cases reported to have gotten tested because of a SwissCovid alert. Of the 185 exposed contacts, 132 (71%) used the app. Of those 132 app users, 46 (35%) received the SwissCovid warning related to the exposure that brought them into contact with the cantonal authorities.

## Evidence for app users responding to notifications

Routine public health surveillance data suggest that notified contacts seek SARS-CoV-2 testing (Figure 1, Box 4). Information on reason for RT-PCR testing has been collected during the study period and was available for 7,842 (63%) of the 12,456 confirmed cases. Among these, 6,380 reported symptoms compatible with COVID-19, 487 reported outbreak investigation, and 41 reported the SwissCovid app as the reason for the test (Figure 1, Box 5). As the information on the reason for the test was only complete for 63% of confirmed cases, the total number of cases that were tested because of the notification by the app is likely higher. To estimate this number, we applied multiple imputation by chained equation [18], accounting for the effect of age and sex on the probability of missing information. This approach yielded a total of 65 cases (95% CI: 54-77) reporting the SwissCovid app as the reason for testing over the period considered (Figure 2). These cases displayed a slightly younger age distribution but a similar sex distribution compared to cases reporting another reason for testing (Figure 3C).

## Effectiveness of the SwissCovid app

A key measure to quantify the effectiveness of contact tracing at identifying SARS-CoV-2 infections is the number of positive contacts per index case. This number depends on several factors, such as the number of contacts traced per index case and the overall dynamics of the epidemic. Two large studies of classic contact tracing for SARS-CoV-2 found 23 secondary cases in contacts of 100 index cases (0.23, 95% CI: 0.15-0.32) in Taiwan [19] and 2,169 positive cases in contacts of 5,706 index cases (0.38, 95% CI: 0.37-0.39) in South Korea [20]. To compare the effectiveness of the SwissCovid app to these classic contact tracing studies, we estimated the number of notified positive contacts using the app per index case who entered a Covidcode using the formula ε=n/(cμ), where n=65 (95% CI: 54-77) is the imputed total number of confirmed cases that reported the SwissCovid app as the reason for the test, c=1,645 is the number of entered Covidcodes, and μ=16.7% is the proportion of the Swiss population that are active users of the app, weighted by the number of confirmed cases per day. Hence, the term cμ corresponds to the number of index cases entering Covidcodes scaled by the probability that their contacts use the SwissCovid app assuming a homogeneous distribution of the app coverage. We obtained ε=0.24 (95% CI: 0.20-0.27), which is in a similar range to the numbers from the classic contact tracing studies.

## DISCUSSION

Several factors could affect the estimated effectiveness of the SwissCovid app. Due to clustering of app users, the calculated ε could represent an upper estimate as the uptake of the SwissCovid app in contacts of app users might be higher than the average uptake in the Swiss population. Assuming μ=71%, which is the uptake of the app in contacts of RT-PCR-confirmed cases from the Zurich SARS-CoV-2 Cohort, we obtained ε=0.06 (95% CI: 0.05-0.06) as a lower estimate which would still represent a respectable effectiveness of the app at identifying SARS-CoV-2 infections in contacts of index cases. However, a number of factors can also contribute to an underestimate of ε. First, confirmed cases might only report the presence of symptoms as the reason for the test even though they were notified by the app. Second, due to the decentralized and voluntary nature of SwissCovid, there may have been more notified contacts that got infected and self-isolated following the notification, but did not get tested, and are thus missing from the analysis. Third, reported numbers of RT-PCR-confirmed cases using the app for the study period might be slightly higher in reality due to time delays in reporting.

Reliable, continuous monitoring of app effectiveness indicators is still being improved. Also, citizen reports on social media (e.g. Twitter) have suggested multi-day delays in receiving Covidcodes following positive test results in some instances. Along the same lines, during the study period, only about two thirds of issued Covidcodes have been entered and triggered notifications (Figure 3B). Efforts to streamline procedures and app user interactions with local health authorities - i.e. a direct referral of notified hotline callers to the responsible local authorities - are underway. Our findings illustrate that digital contact tracing can be effective even with low uptake, as suggested by mathematical modeling [21]. Because app coverage affects both the number of index cases and their contacts, the total number of SARS-CoV-2 infections that could be identified through digital contact tracing scales with the square of the coverage and could substantially increase with higher uptake (Figure 3D).

In conclusion, based on the data collected during the initial deployment of the SwissCovid app, we argue that voluntary digital contact tracing can show similar effectiveness at identifying infected partners of index cases as classic contact tracing, provided that both the index case and the exposed contacts use the app. As the effectiveness of digital contact tracing crucially depends on a strong embedding into an efficient testing and contact tracing infrastructure on the ground, apps like SwissCovid represent a helpful complementary tool for controlling the spread of SARS-CoV-2. The strength of evidence for app effectiveness as summarized in Figure 1 illustrates that most indicators have reached at least a “proof-of-principle” stage. That is, the outcome of interest was observed independently in two data sources in all important key indicators. There is, however, still room for improvement. Upcoming improvements in the EN API by Apple and Google will increase the precision in determining risk, and reduce delay in communicating it to users. International interoperability exchanges, planned for October 2020, will increase the effectiveness of the app, in particular in the countries bordering Switzerland. Speed is essential to the effectiveness of TTIQ strategies [4,5]. Reducing the time to quarantine for contacts, as a result of digital contact tracing, should provide an additional, important benefit to COVID-19 mitigation efforts.

## Data Availability

Data to reproduce results will be available upon peer-reviewed publication.

## ACKNOWLEDGEMENT

The authors would like to acknowledge the Swiss Federal Office of Public Health, the Federal Statistical Office, and the Federal Office of Information Technology, Systems and Telecommunication for their collaborative contributions which were essential for the development, deployment, and assessment of SwissCovid. We thank the epidemiology unit at the Federal Office of Public Health for the contributions in providing data and commenting on the results and conclusions. We would also like to thank the Cantonal Health Directorate Zurich for their support and collaboration in the conduct of the Zurich SARS-CoV-2 Cohort, as well as the Swiss School of Public Health and the Corona Immunitas program for contributing to the study with their structure and services.

## FUNDING

MS received funding from the European Union’s Horizon 2020 research and innovation programme - project “Versatile emerging infectious disease observatory - forecasting, nowcasting and tracking in a changing world (VEO)” (No. 874735). CA received funding from the European Union’s Horizon 2020 research and innovation programme - project EpiPose (No 101003688) and the Swiss National Science Foundation (grant 196046). NL received funding from the European Union’s Horizon 2020 research and innovation programme - project EpiPose (No 101003688) and the Swiss National Science Foundation (grant 176233). TB, DM and VvW received funding from the Cantonal Health Directorate Zurich, the Federal Office of Public Health, the University of Zurich Foundation Pandemic Fund and the Fondation Les Mûrons for the Zurich SARS-CoV-2 Cohort.

## Competing Interests

The authors declare that there are no competing interests.

## Author Contribution

All authors contributed to the writing of the paper. CA, NA, JR, VW, DM, and TB analyzed the data.

## Notes

### Competing Interest Statement

The authors have declared no competing interest.

### Author Declarations

The Zurich Sars-CoV-2 Cohort Study has been approved by the Ethics Commission of the Canton of Zurich (BASEC Nr. 2020-01739). Written informed consent has been obtained from all participants.

### Summary of Updates

Updated with new figure, some clarifications

## REFERENCES

1. World Health Organization. Coronavirus disease (COVID-19) - Weekly Epidemiological Update. [cited 4 Sep 2020]. Available: https://www.who.int/docs/default-source/coronaviruse/situation-reports/20200831-weekly-epi-update-3.pdf?sfvrsn=d7032a2a_4

2. Salathé M, Althaus CL, Neher R, Stringhini S, Hodcroft E, Fellay J, et al. COVID-19 epidemic in Switzerland: on the importance of testing, contact tracing and isolation. Swiss Medical Weekly. 2020. doi:https://doi.org/10.4414/smw.2020.20225

3. Aleta A, Martin-Corral D, Piontti AP y, Ajelli M, Litvinova M, Chinazzi M, et al. Modeling the impact of social distancing, testing, contact tracing and household quarantine on second-wave scenarios of the COVID-19 epidemic. Medrxiv. 2020; 2020.05.06.20092841. doi:10.1101/2020.05.06.20092841

4. Kucharski AJ, Klepac P, Conlan AJK, Kissler SM, Tang ML, Fry H, et al. Effectiveness of isolation, testing, contact tracing, and physical distancing on reducing transmission of SARS-CoV-2 in different settings: a mathematical modelling study. Lancet Infect Dis. 2020. doi:10.1016/s1473-3099(20)30457-6

5. Ferretti L, Wymant C, Kendall M, Zhao L, Nurtay A, Abeler-Dörner L, et al. Quantifying SARS-CoV-2 transmission suggests epidemic control with digital contact tracing. Science. 2020;368: eabb6936. doi:10.1126/science.abb6936

6. Troncoso C, Payer M, Hubaux J-P, Salathé M, Larus J, Bugnion E, et al. Decentralized Privacy-Preserving Proximity Tracing. arxiv. 2020. Available: https://arxiv.org/abs/2005.12273

7. Wyl V von, Bonhoeffer S, Bugnion E, Salathé M, Stadler T, Troncoso C, et al. A research agenda for digital proximity tracing apps. Swiss Medical Weekly. 2020. doi: https://doi.org/10.4414/smw.2020.20324

8. World Health Organization. Ethical considerations to guide the use of digital proximity tracking technologies for COVID-19 contact tracing. [cited 4 Sep 2020]. Available: https://www.who.int/publications/i/item/WHO-2019-nCoV-Ethics_Contact_tracing_apps-2020.1

9. Gasser U, Ienca M, Scheibner J, Sleigh J, Vayena E. Digital tools against COVID-19: taxonomy, ethical challenges, and navigation aid. Lancet Digital Heal. 2020;2: e425–e434. doi:10.1016/s2589-7500(20)30137-0

10. Telecommunication SFO of ITS and. SwissCovid Exposure Score Calculation. [cited 4 Sep 2020]. Available: https://github.com/admin-ch/PT-System-Documents/blob/master/SwissCovid-ExposureScore.pdf

11. World Health Organization. Contact tracing in the context of COVID-19. [cited 4 Sep 2020]. Available: https://www.who.int/publications/i/item/contact-tracing-in-the-context-of-covid-19

12. Wyl V von, Hoeglinger M, Sieber C, Kaufmann M, Moser A, Serra-Burriel M, et al. Are COVID-19 proximity tracing apps working under real-world conditions? Indicator development and assessment of drivers for app (non-)use. medRxiv. 2020. doi:10.1101/2020.08.29.20184382

13. Swiss Federal Office of Statistics. Calculation methods for estimating the number of active SwissCovid apps. [cited 4 Sep 2020]. Available: https://www.experimental.bfs.admin.ch/bfsstatic/dam/assets/13667538/master

14. Swiss Federal Office of Statistics. SwissCovid App Monitoring. [cited 4 Sep 2020]. Available: https://www.experimental.bfs.admin.ch/expstat/en/home/innovative-methods/swisscovid-app-monitoring.html

15. Corona Immunitas. [cited 4 Sep 2020]. Available: https://www.corona-immunitas.ch/program

16. Stringhini S, Wisniak A, Piumatti G, Azman AS, Lauer SA, Baysson H, et al. Seroprevalence of anti-SARS-CoV-2 IgG antibodies in Geneva, Switzerland (SEROCoV-POP): a population-based study. Lancet. 2020;396: 313–319. doi:10.1016/s0140-6736(20)31304-0

17. Zurich Coronavirus Cohort: an observational study to determine long-term clinical outcomes and immune responses after coronavirus infection (COVID-19), assess the influence of virus genetics, and examine the spread of the coronavirus in the population of the Canton of Zurich, Switzerland. [cited 4 Sep 2020]. Available: http://www.isrctn.com/ISRCTN14990068

18. White IR, Royston P, Wood AM. Multiple imputation using chained equations: Issues and guidance for practice. Stat Med. 2011;30: 377–399. doi:10.1002/sim.4067

19. Cheng H-Y, Jian S-W, Liu D-P, Ng T-C, Huang W-T, Lin H-H, et al. Contact Tracing Assessment of COVID-19 Transmission Dynamics in Taiwan and Risk at Different Exposure Periods Before and After Symptom Onset. Jama Intern Med. 2020;180. doi:10.1001/jamainternmed.2020.2020

20. Park YJ, Choe YJ, Park O, Park SY, Kim Y-M, Kim J, et al. Contact Tracing during Coronavirus Disease Outbreak, South Korea, 2020. Emerging Infectous Diseases. 2020. doi:DOI: 10.3201/eid2610.201315

21. Abueg M, Hinch R, Wu N, Liu L, Probert WJM, Wu A, et al. Modeling the combined effect of digital exposure notification and non-pharmaceutical interventions on the COVID-19 epidemic in Washington state. medRxiv. 2020. doi:10.1101/2020.08.29.20184135

